# Field performance of NowCheck rapid antigen test for SARS-CoV-2 in Kisumu County, western Kenya

**DOI:** 10.1101/2021.08.12.21261462

**Authors:** S. N. Onsongo, K. Otieno, S. van Duijn, E. Adams, M. Omollo, I. A. Odero, A. K’Oloo, N. Houben, E. Milimo, R. Aroka, H.C. Barsosio, F. Oluoch, A. Odhiambo, S. Kariuki, T.F. Rinke de Wit

## Abstract

**Background:** Low- and middle-income countries (LMICs) are increasingly adopting low-cost Coronavirus disease 2019 (COVID-19) rapid antigen tests to meet the high demand for SARS-CoV-2 testing. Whilst testing using real-time polymerase chain reaction (RT-PCR) is the current gold standard, its widespread use in LMICs is limited by high costs, turnaround times and is not readily available in most places. COVID-19 antigen tests (Ag-RDT) provide a suitable alternative due to their low cost, rapid turnaround time and easy to set up and use. This study aimed to assess the field performance of the NowCheck COVID-19 antigen kit (Ag-RDT) as a point of care test (POCT) in select healthcare facilities in western Kenya.

**Methods:** We conducted a prospective multi-facility field evaluation study of the NowCheck COVID-19 rapid antigen test (Ag-RDT) compared to SARS-CoV-2 RT-PCR (RT-PCR). After obtaining informed consent, trained laboratory technicians collected two pairs of oropharyngeal and nasopharyngeal swabs, both antigen and RT-PCR testing, first for Ag-RDT and next for RT-PCR. We performed Ag-RDTs on-site and shared the results with both the study participants and their healthcare providers within 15-30 minutes. We carried out all RT-PCR tests in a central referral laboratory. The turnaround time for RT-PCR results was typically 24-48 hours. We captured the results of both methods using an electronic digital application.

**Findings:** Between December 2020 and March 2021, we enrolled 997 participants who met the Kenyan Ministry of Health COVID-19 case definition. The median age of study participants was 39 years (range one to 80 years), with 54% male. Ag-RDT had a sensitivity of 84.5% (76.0-90.8) and a specificity of 94.4% (95% CI: 92.7-95.8) with an accuracy of 94.2% (92.5-95.6) when a cycle threshold value (Ct value) of ≤35 was used. The highest sensitivity of 87.7% (77.2-94.5) was observed in samples with Ct values ≤ 30 and the highest specificity of 97.5% (96.2-98.5) at Ct value of <40.

**Interpretation:** The NowCheck COVID-19 Ag-RDT showed good performance in field evaluation in multiple healthcare facilities in a developing country. The sensitivity of the kit exceeded the minimum recommended cut-off of 80% as recommended by WHO^1^. The high specificity of this kit at 94.4% at Ct values ≤33 and 97.5% at Ct values <40 matched that of real-time PCR, making it a good rule-out test for symptomatic patients with COVID-19-like symptoms. The faster turnaround time to results, lower cost, simple analytical steps requiring no equipment or infrastructure makes antigen testing an attractive field-screening method to meet the high demand for COVID-19 testing.

**Funding:** Achmea Foundation, Pfizer Foundation, Dimagi and the Netherlands Ministry of Foreign Affairs supported this project. The funding sources did not have any role in study design, data collection, analysis, interpretation, summarizing the data or decision to submit the manuscript for publication.

## Background

Since it was first described in December 2019 in Wuhan, Hubei Province in China, the severe acute respiratory syndrome coronavirus 2 (SARS-CoV-2) causes coronavirus disease 2019 (COVID-19) has caused a significant global public health problem^2^. As of 27 July 2021, SARS-COV-2 had resulted in 195,1963,640 infections resulting in 4,180,182 deaths in over 220 countries^3^. Preventing the spread of SARS-COV-2 is a global public health priority requiring multiple interventions such as social distancing, hand hygiene, infection protection devices, vaccination, timely and accurate diagnosis, and treatment of the sick ^4,5^.

Reverse transcriptase-polymerase chain reaction (RT-PCR) of nasal/oropharyngeal samples is the gold standard diagnostic method of SARS-CoV-2^6^. Though RT-PCR has high sensitivity, it requires a robust infrastructure, well-trained staff, an elaborate supply chain to support testing services, and it comes at a high cost ^7^. In many LMICs, there are significant gaps to support RT-PCR testing, resulting in limited access to testing. Although a typical RT-PCR assay takes an average of 4-6 hours, in most countries, the time to get results is much longer due to logistics, inefficient reporting systems and lack of digital data management ^8,9^. Furthermore, access is much more limited in hard-to-reach communities, making controlling the pandemic a problematic task. With low vaccination rates not exceeding 2% in Africa and across many low-and middle-income countries, testing remains a crucial pillar in the fight against SARS-COV-2^10,11^. In these settings, point of care Ag-RDTs provides a practical alternative to RT-PCR. Although Ag-RDTs may not fully replace RT-PCR testing, they offer an easy-to-use POCT method. Moreover, Ag-RDTs can easily be scaled to community settings since they do not require any equipment or well-trained laboratory staff. The quick turnaround time of 15-30 minutes helps in faster triage, rapid mass testing, correct patient placement, and, where applicable, support contact tracing^12^. The low cost per test and ease of use of Ag-RDTs makes them a suitable public health tool to widely scale-up testing in LMICs even in remote communities and in high transmission zones^12^.

The World Health Organization (WHO) recommends using antigen test kits with a sensitivity of at least 80% and specificity of greater than 97% compared to the gold standard RT-PCR ^13^. In these settings, such Ag-RDTs provides a decent alternative to RT-PCR, hence the rapid increase in production of several such kits in the market within a short period. As of 25 July 2021, less than 20 months since the COVID-19 pandemic started, 176 listed antigen kits on the Foundation for Innovative New Diagnostics, (a global alliance for diagnostics that seeks to prioritize, partner, develop, evaluate, and support the equitable implementation of diagnostic tests) website^14, 15^.

Although various kit manufacturers report the performance of their diagnostic kits, their operational performance can vary when subjected to the field or real-world conditions. Diagnostic performances of most Ag-RDTs in the market have yet to be adequately evaluated in field situations in LMICs. Moreover, most regulators carry out only limited, laboratory-based Ag-RDT evaluations before granting emergency use authorizations for use^16^. This study aimed at an in-depth evaluation of the field performance of BioNote NowCheck COVID-19 Ag-RDT among Kenyans suspected to have COVID-19 attending four health facilities in Kisumu County, western Kenya.

## Materials and Methods

### Study design and oversight

We conducted a cross-sectional, prospective diagnostic evaluation study comparing the field-based operational performance of the NowCheck SARS-CoV-2 Ag-RDT against the gold standard RT-PCR. Four health facilities in Kisumu County participated, consisting of private hospitals, public hospitals, and faith-based hospitals, all of which were part of a public-private partnership in response to the SARS-CoV-2 pandemic in Kisumu dubbed “ COVID-Dx”. We obtained research and ethical approval from Jaramogi Oginga Odinga Teaching & Referral Hospital (JOOTRH) under IERC/JOOTRH/334/20 and Research License from National Commission for Science, Technology, and Innovation, license number BAHAMAS ABS/P/20/7959. We obtained informed consent for all eligible adults and assent and parental consent for eligible minors below 18 years. All study procedures were carried out according to ethical principles of the Helsinki Declaration of 1964 and good clinical practices. All study personnel were appropriately trained on infection prevention control using the Kenyan Ministry of Health guidelines and provided with personal protective equipment.

### Sample collection

Trained laboratory technicians collected paired nasopharyngeal and oropharyngeal samples from eligible participants attending four healthcare facilities in Kisumu County. Eligible participants met the Ministry of Health COVID-19 case definition and provided informed consent before sample collection. Pre- and post-test counselling services were provided as additional support for participants.

### Antigen testing

The NowCheck SARS-CoV-2 Ag test is a rapid antigen chromatographic immunoassay for the qualitative detection of N protein of SARS-CoV-2 antigens^17^. Trained health workers carried out the testing procedure according to the manufacturer’s specifications. The results were provided to patients within 15-30 minutes of testing while awaiting final RT-PCR results within 24-48 hours. The results were digitally captured onto a tablet/smartphone application (digitized Kenyan COVID-19 Case Investigation Form) and stored in a safe cloud server as described here^18^, together with general information on the patients (such as age, gender and occupation). Results release, patient care and follow up was performed according to the Kenya Ministry of Health guidelines^19^.

### Reverse transcription-polymerase chain reaction (RT-PCR)

RT-PCR testing was centrally performed at Kenya Medical Research Institute (KEMRI). SARS-CoV-2 viral ribonucleic acid (RNA) extraction from the paired nasopharyngeal and oropharyngeal samples was manually done using MagMAX™ Viral RNA Isolation Kit (Thermo Fisher Scientific, MA, USA). Post-RNA extraction, real-time SARS-CoV-2 PCR was carried out using TaqPath™ 19 kit (Thermo Fisher Scientific, MA, USA). Three primer sets specific to different SARS-CoV-2 genomic regions (ORF1ab, N gene, S gene) were used for the RT-PCR reaction. Probes for bacteriophage MS2 were added as an RNA control to verify the efficacy of the sample preparation and the absence of inhibitors in the RT-PCR reaction. The assays were set up on the 7500 Fast Real-Time PCR Instrument (Applied Biosystems, Thermo Fisher Scientific, MA, USA). A 5 µl template RNA from each sample’s nucleic acid was used with 20 µl of the PCR master-mix.

The following thermocycling conditions were employed; 2 min at 25°C incubation, 10 min at 53°C for reverse transcription, 2 min at 95 °C for enzyme activation and 40 cycles of 3 s at 95°C and 30s at 60°C. Samples having exponential growth curve and Ct < 40 in at least two SARS-CoV-2 targets were considered positive, while those with Ct values in only one target were inconclusive. Assays with all targets, including MS2 negative, were deemed invalid. All invalid and inconclusive assays were repeated. All test procedures were performed according to manufacturer-prescribed protocols. The antigen test results were compared with gold-standard RT-PCR results and used to compute kit field performance specifications. Table 1 below provides the breakdown of paired antigen and PCR testing across the four facilities marked A to D.

**Table 1:** Demographic characteristics of study participants

| Variable | n (%) | 95%CI |
| --- | --- | --- |
| **Sex** |  |  |
| *Male* | 536(53.8) |  |
| *Female* | 461(46.2) |  |
| **Occupation** |  |  |
| *Informal workers* | 367(36.8) | 33.86-39.86 |
| *Formal workers (non-healthcare)* | 276(27.7) | 24.98-30.55 |
| *Students* | 163(16.4) | 14.17-18.78 |
| *Healthcare workers* | 124(12.4) | 10.52-14.64 |
| *Unemployed* | 42(4.2) | 3.12-5.65 |
| *Children (<14 yrs)* | 25(2.5) | 1.69-3.69 |
| **Symptoms** |  |  |
| *Cough* | 738(16.1) | 15.08-17.22 |
| *Headache* | 598(13.1) | 12.12-14.07 |
| *Sore throat* | 596(13.0) | 12.07-14.03 |
| *General weakness* | 548(12.0) | 11.06-12.94 |
| *Nausea/vomiting* | 508(11.1) | 10.22-12.04 |
| *History of fever/chills* | 426(9.3) | 8.50-10.18 |
| *Runny nose* | 409(8.9) | 8.14-9.80 |
| *Diarrhoea* | 271(5.9) | 5.27-6.64 |
| *Pain-muscular/abdominal/chest/joint* | 253(5.5) | 4.90-6.22 |
| *Irritability/confusion* | 138(3.0) | 2.55-3.55 |
| *Shortness of breath* | 80(1.8) | 1.40-2.17 |
| *Other* | 8(0.2) | 0.08-0.34 |
| *Asymptomatic* | 3(0.1) | 0.02-0.20 |

**Table 2:** Distribution of antigen and PCR testing across facilities in Kisumu County.

| Facility | Ag-RDT results per Facility |  |  | SARS CoV 2 RT-PCR |  |  |
| --- | --- | --- | --- | --- | --- | --- |
|  | Negative | Positive | Total RDTs | Negative | Positive | Total RT-PCR |
| Facility A | 106 | 45 | 151 | 104 | 47 | 151 |
| Facility B | 644 | 72 | 716 | 627 | 89 | 716 |
| Facility C | 71 | 10 | 81 | 71 | 10 | 81 |
| Facility D | 47 | 2 | 49 | 43 | 6 | 49 |
| Totals | 868 | 129 | 997 | 845 | 152 | 997 |

### Statistical analysis

Analyses were performed using STATA-15 (StataCorp LLC, Texas, USA) and Microsoft Excel (Microsoft, Redmond, Washington, USA). We used descriptive statistics to describe the general information of study participants. Continuous data were summarized and presented in means, standard deviation (SD), median, and range as applicable. Categorical data were summarized in percentages, and a significance value of 0.05 in statistic tests was considered. Ag-RDT sensitivity, specificity, positive likelihood ratio, negative likelihood ratio, positive predictive value, negative predictive value, and accuracy were calculated using MedCalc online software calculator^20^, grouped according to Ct values using the following cut-offs: 25, 30, 35 and 40

## Results

### Demographic characteristics

From 28 December 2020 and 31 March 2021, 997 eligible participants were enrolled in the study across four health facilities. The median age was 37 years (interquartile range, 28–49 years). Their demographic characteristics are summarized in Table 1. The distribution of antigen and RT-PCR testing across the four participating health facilities in Kisumu County are summarized in Table 1 below.

### Performance characteristics

When compared against RT-PCR as the gold standard, the kit sensitivity and specificity showed variable performance across varying Ct values (Table 3). The lowest sensitivity, 71.5% (63.2-78.6), was reported at Ct value <40, while the lowest specificity of 89.5% (87.4-91.4) was noted at the lowest Ct values. We observed the best Ag-RDT performance when the Ct values were ≤35 with a sensitivity of 84.5% (76.00% to 90.9) and specificity of 95.3% (93.7-96.6). At this cut-off, the diagnostic accuracy was 94.2% (92.5-95.6). The highest sensitivity of 87.7% (77.2-94.5) was observed in samples with Ct values ≤30, corresponding with samples with higher viral loads. At Ct value <40, kit sensitivity dropped to 71.5% (63.2-78.6) with a specificity matching that of RT-PCT at 97.5% (96.2-98.5).

**Table 3:** Performance characteristics of NowCheck COVID-19 Ag-RDT across grouped PCR Ct values.

| | Ct $\leq 25$ | Ct $\leq 30$ | Ct $\leq 35$ | Ct $\leq 40$ |
| --- | --- | --- | --- | --- |
| Sensitivity | 87.5% (71.0-96.5) | 87.7% (77.2-94.5) | 84.5% (76.0 - 90.85) | 71.5% (63.2-78.6) |
| Specificity | 89.5% (87.4-91.4) | 92.3% (90.4-93.9) | 95.30% (93.7 - 96.59) | 97.5% (96.2-98.5) |
| Positive Likelihood Ratio | 8.4(6.7-10.5) | 11.4(8.9-14.4) | 17.98 (13.2 to 24.4) | 28.81(18.7-44.5) |
| Negative Likelihood Ratio | 0.14(0.1-0.4) | 0.13(0.1-0.3) | 0.16 (0.1 - 0.3) | 0.29(0.2-0.4) |
| Positive Predictive Value | 21.7% (18.1-25.8) | 44.2% (38.4-50.2) | 67.4% (60.4 -73.79) | 83.7% (76.9-88.1) |
| Negative Predictive Value | 99.5% (98.9-99.8) | 99.1% (98.3-99.5) | 98.16% (97.1% - 98.8) | 95.1% (93.7-96.1) |
| Accuracy | 89.5% (87.5-91.3) | 91.9% (90.1-93.6) | 94.18% (92.5 to 95.6) | 93.6% (91.9-95.0) |

## Discussion

Testing is essential for controlling SARS-COV-2 infection, allowing early identification and isolation of cases to slow down transmission, and providing timely clinical management to those affected and protecting health systems operations through triaging at admissions^21^. In most LMICs, access to RT-PCR testing capacity is limited to a few specialized centres resulting in significant delays in testing ^22,23^. Ag-RDTs presents a suitable testing method with several advantages. These include low-cost, availability as point-of-care, easy to use, do not require electricity, specialized skills or special equipment. The lower cost per test and ease of rolling out to remote communities may offer opportunities to rapidly scale up to meet the high demand for testing, contributing to effective control of the pandemic^13,24^.

In its preliminary guidance on using SARS-CoV-2 rapid antigen kits, the WHO recommends using kits with a sensitivity of ≥ 80% and specificity greater than 97%^13^. In this study, we sought to evaluate the field performance of the NowCheck SARS-CoV-2 antigen kit in a field context, comparing it with the manufacturer’s reported sensitivity and specificity of 89.2% and 97.3%, respectively. In this field evaluation, the BioNote NowCheck kit shows an excellent sensitivity of 84.5% (76.0% to 90.7%) and specificity 95.30% (93.7% to 96.6%) when Ct value ≤35 was used. The sensitivity dropped to 71.5% (63.2-78.6) and specificity increased to 97.5% (96.2-98.5) when using a Ct value of ≤40. The highest sensitivity was in samples with a Ct value ≤ 30, corresponding to a higher viral load. At Ct values ≤ 30, the sensitivity was 87.7% (77.2-94.5) and specificity at 92.3% (90.4-93.9).

Studies from around the world evaluating various rapid diagnostic antigen kits show mixed findings ^25^. Reported sensitivities vary from as low as 30.2% for Coris COVID-19 Ag Respi-Strip^26^ to 64% for the Alltest lateral flow immunoassay ^27^ to kits with reported sensitivities higher than 80% such as Orient Gene, Deepblue, Abbott and Innova SARS-CoV-2 Antigen Rapid^28^. In a field evaluation study by Eliseo et al., similar to our study in a primary health centre, the Panbio Ag-RDT kit showed a sensitivity of 79.6% and specificity of 100%^29^. In a similar field evaluation of Standard Q Ag-RDT in Uganda, Nalumansi et al.^30^ noted a less than optimal performance with a sensitivity of 70% and specificity of 92%. The authors noted better performance at lower Ct values ≤29 with sensitivity exceeding 92%. The same finding was confirmed by Nahal et al.^31^, who evaluated seven different rapid antigen kits and noted better performance at higher viral load, with an optimal Ct value of ≤25.

To our knowledge, this is the first study in Kenya to extensively evaluate the field performance of an affordable COVID-19 Ag-RDT kit. The Ag-RDT kit showed excellent sensitivity and specificity compared to the gold standard, despite the challenges of the real-world environment. Such challenges included variability in sample collection practices among laboratory technicians across different healthcare facilities. Rapid turnover of healthcare staff required regular retraining. Growing stigma and fear due to curfews, lockdowns and stringent quarantine measures contributed to lab staff hesitation to collect samples (to the level of requesting ‘danger allowances’). Shipment of samples to the central lab through motorbikes experienced cooling challenges. Patients became increasingly critical to getting tested, given the limited perspectives offered when testing positive. Stock-outs of reagents and kits for RT-PCR occurred, as well as (temporary) breakdowns of crucial lab equipment.

In general, the Ag-RDT kit proved easy to use, requiring limited training time. Patients’ discomfort during sample taking was reportedly minimal, and they appreciated an earlier (preliminary) COVID-19 test result. Patients and health workers considered the immediate availability of pre-and post-test counselling services of good value in nudging subsequent patient behaviour, including quarantining, support of contact tracing and hospitalization of those in need. This study provides a proof of concept that rapid antigen kits can easily be rolled out to the community level to support the quick point-of-care diagnosis that allows rapid patient triage, quarantining and hospital placement. In areas with limited access to RT-PCR testing and high patient flow, Ag-RDTs can be scaled up to remote locations, support quick turnaround times for test results with minimal training requirements. Rapid testing can help quick and effective contact tracing at the point of care, supporting efforts to contain SARS CoV-2 spread within the community.

Though helpful, the limitations of these kits should be noted by the users. Some studies have suggested low sensitivity in screening asymptomatic patients ^29,31,32^. Various authors have cited lower sensitivity in patients with low viral load (high Ct values on RT-PCR); hence RT-PCR testing should continue to be offered where clinical suspicion is high^24,30,33^. The users should follow manufacturers laid down testing procedures to avoid potentially misleading false-negative or false-positive results.

## Data Availability

All data relating to this manuscript is available

## Acknowledgements

The Netherlands Ministry of Foreign Affairs supported this work through a core grant to Health Insurance Fund Foundation, which supports PharmAccess Foundation and by complementary grants from Achmea Foundation and Pfizer Foundation. The CommCare mobile data collection software was provided by Dimagi free of charge. BioNote Inc provided free RDTs for evaluation. Many thanks to all staff who supported this project.

## Notes

### Competing Interest Statement

The authors have declared no competing interest.

### Author Declarations

Research and ethical approval from Jaramogi Oginga Odinga Teaching & Referral Hospital (JOOTRH) under IERC.IB/VOL.II/355/20, and Research License from National Commission for Science, Technology, and Innovation, license number ABS/P/20/7959.

